# Deep learning predicts post-surgical recurrence of hepatocellular carcinoma from digital whole-slide images

**DOI:** 10.1101/2020.08.22.20179952

**Authors:** Rikiya Yamashita, Jin Long, Atif Saleem, Daniel L. Rubin, Jeanne Shen

## Abstract

Recurrence risk stratification of patients undergoing primary surgical resection for hepatocellular carcinoma (HCC) is an area of active investigation, and several staging systems have been proposed to optimize treatment strategies. However, as many as 70% of patients still have tumor recurrence at 5 years post-surgery. Routine hematoxylin and eosin (H&E)-stained histopathology slides may contain morphologic features associated with tumor recurrence. In this study, we developed and independently validated a deep learning-based system (HCC-SurvNet) that provides risk scores for disease recurrence after primary surgical resection, directly from H&E-stained digital whole-slide images of formalin-fixed, paraffin embedded liver resections. Our model achieved a concordance index of 0.724 on a held-out internal test set of 53 patients, and 0.683 on an external test set of 198 patients, exceeding the performance of standard staging using the American Joint Committee on Cancer (AJCC)/International Union against Cancer (UICC) Tumor-Node-Metastasis (TNM) classification system, on both the internal and external test cohorts (p = 0.018 and 0.025, respectively). We observed statistically significant differences in the survival distributions between low- and high-risk subgroups, as stratified by the risk scores predicted by HCC-SurvNet on both the internal and external test sets (log-rank p-value: 0.0013 and < 0.0001, respectively). On multivariable Cox proportional hazards analysis, the risk score was an independent risk factor for post-surgical recurrence, on both the internal (hazard ratio (HR) = 7.44 (95% CI: 1.60, 34.6), p = 0.0105) and external (HR = 2.37 (95% CI: 1.27, 4.43), p = 0.0069) test sets. Our results suggest that deep learning-based models can provide recurrence risk scores which may augment current patient stratification methods, and help refine the clinical management of patients undergoing primary surgical resection for HCC.

## Introduction

Hepatocellular carcinoma (HCC) is the most prevalent primary liver malignancy and the fourth leading cause of cancer-related death worldwide.^1,2^ Despite advances in prevention, surveillance, early detection, and treatment, its incidence and cancer-specific mortality continue to rise, with the majority of patients still presenting at advanced stages.^1,2^ To stratify patients according to their expected outcome in order to optimize treatment strategies, several staging systems, such as the American Joint Committee on Cancer (AJCC)/International Union against Cancer (UICC) Tumor-Node-Metastasis (TNM)^3^ and the Barcelona Clinic Liver Cancer (BCLC) system,^4^ have been proposed and validated. However, as many as 70% of patients still have tumor recurrence within 5 years post-treatment,^2,5–7^ including both true recurrence due to intrahepatic metastasis and de novo primary cancers arising in the background liver, as the majority of HCCs occur in patients with underlying chronic liver disease that directly contributes to the development of HCC. Therefore, further refinement and improvement of recurrence risk stratification is warranted.

Histopathologic assessment plays a key role in recurrence risk stratification, as it evaluates human-recognizable morphologic features associated with tumor recurrence, such as histopathologic grade and vascular invasion.^8–11^ Prognostic nomograms for prediction of recurrence after curative liver resection for HCC have been proposed using clinicopathologic variables.^12^ However, histopathologic features are interpreted by pathologists, which is subject to reproducibility problems (an example being inter- and intra-observer variability in the assessment of microvascular invasion^13^). On the other hand, recent advances in computer vision, deep learning, and other forms of machine learning have enabled the identification of histomorphologic patterns and features informative of disease outcomes which are not readily recognizable by the human eye, and which are reproducible. Thus, there has been much interest in applying computer vision methods to histologic images for automated outcome prediction.^14–21^ Mobadersany et al.^14^ and Zhu et al.^15^ applied convolutional neural networks, a type of deep learning network, to predict patient survival directly from histopathologic images of brain and lung cancers, respectively. In these two studies, to achieve direct survival prediction from histopathologic images, the negative partial log-likelihood was used as the loss function, which enabled the models to output the risk values of the Cox proportional hazard model’s exponential part. Saillard et al.^21^ recently developed a deep learning-based model for the prediction of overall survival after surgical resection in patients with HCC, using digital whole-slide images. However, no studies to date have sought to predict post-surgical recurrence of HCC directly from histopathologic images using deep learning.

In this study, we developed and independently validated a deep convolutional neural network for predicting risk scores for the recurrence-free interval (RFI) after curative-intent surgical resection for HCC, directly from digital whole-slide images (WSI) of hematoxylin and eosin (H&E)-stained, formalin-fixed, paraffin embedded (FFPE) primary liver resections. We built on and extended the aforementioned prior work by applying the negative partial log-likelihood as a loss function, so that the model outputs risk scores for post-surgical recurrence. In doing so, we present a fully automated approach to HCC recurrence risk prognostication on histopathologic images, which can be adopted for use in clinical settings to refine treatment and follow-up plans.

## Results

An overall framework for the deep learning-based system for predicting the risk score for RFI, hereafter referred to as HCC-SurvNet, is shown in Figure 1. The system consists of two stages, i.e. tumor tile classification and risk score prediction.

**Figure 1:**
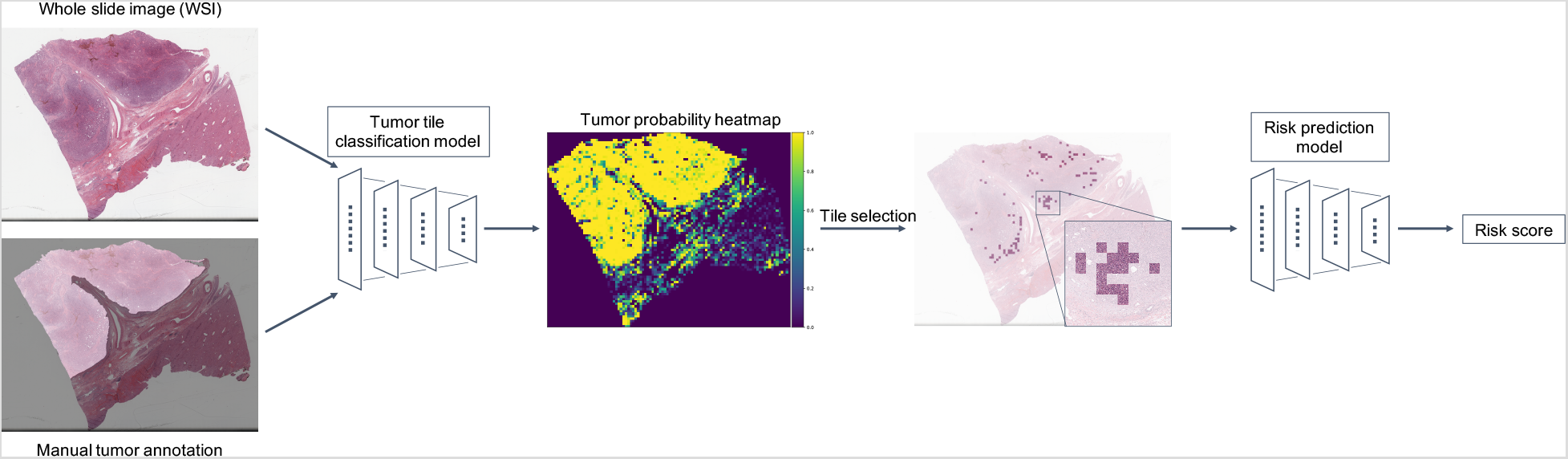
Overview of HCC-SurvNet. All WSI were preprocessed by discarding non tissue-containing white background using thresholding, then partitioned into non-overlapping tiles of size 299 × 299 pixels and colornormalized. A tumor tile classification model was developed using the Stanford-HCCDET dataset, containing WSI with all tumor regions manually annotated. The tumor tile classification model was subsequently applied to each tissue-containing image tile in the TCGA-HCC (n = 360 WSIs) and Stanford-HCC (n = 198 WSIs) datasets for inference. The 100 tiles with the highest predicted probabilities of being tumor tiles were input into the downstream risk prediction model to yield tile-based risk scores, which were averaged to generate a WSI-level risk score for recurrence. Abbreviations: WSI, whole-slide image

### Tumor tile classification

To develop a deep convolutional neural network (CNN) to automatically detect tumor-containing tiles within WSI, we used the Stanford-HCCDET (n = 128,222 tiles from 36 WSI) dataset. All tumor regions in each WSI in the Stanford-HCCDET dataset were manually annotated by the reference pathologist (J.S.). Each WSI was preprocessed and tiled into image patches. Using these ground truth labels and image tiles, we trained and tested a CNN using 78% of WSI in the Stanford-HCCDET for training, 11% for validation, and 11% for internal testing, with no patient overlap between any of these three sets. The final optimized tumor versus non-tumor tile classifier was externally tested on 30 WSI (n = 82,532 tiles) randomly sampled from the TCGA-HCC dataset.

Among the tiles in the internal test set, 25.7% (2,932 of 11,412 tiles) were tumor positive, whereas 48.8% (40,288 out of 82,532 tiles) were tumor positive in the external test set. The accuracies of tumor tile classification were 92.3% and 90.8% for the internal and external test sets, respectively. The areas under the receiver-operating-characteristic-curve (AUROCs) were 0.952 (95% CI: 0.948, 0.957) and 0.956 (95% CI: 0.955, 0.958) for the internal and external test sets, respectively. Model outputs showed a statistically significant difference between tiles with a ground truth of tumor versus non-tumor, on both the internal and external test sets (p< 0.0001 and p< 0.0001, respectively) (Figures 2 and 3).

**Figure 2:**
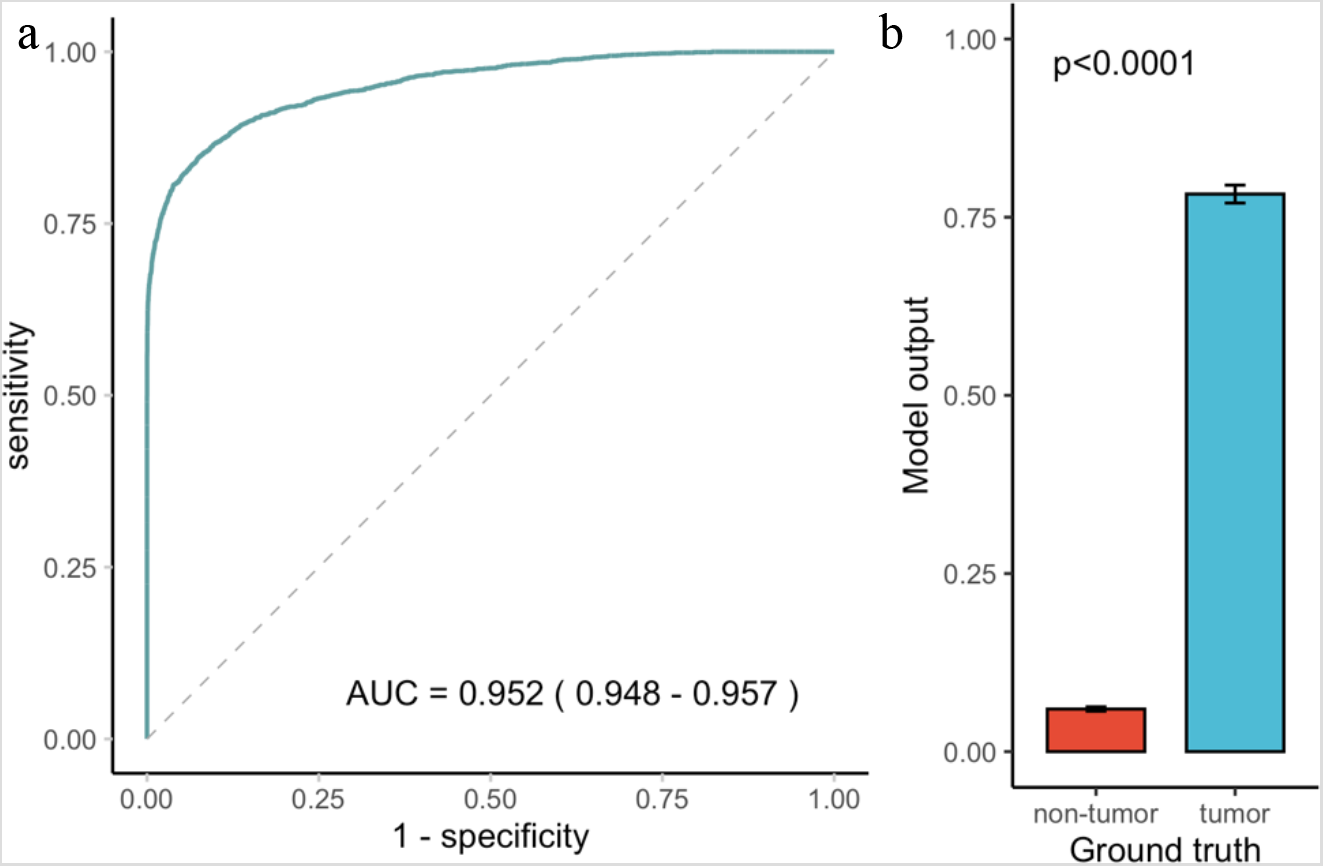
Performance of the tumor tile classification model on the internal test set. The AUROC for tumor tile classification was 0.952 (95% CI: 0.948, 0.957) on the internal test set (a). Model outputs differed significantly between tiles with a ground truth of tumor versus non-tumor (p-value< 0.0001) (b). *The 95% CI for AUC is shown in parentheses in the ROC plot. **Error bars represent 95% CI in the bar chart. The p-value was computed using the Wilcoxon rank sum test. Abbreviations: AUC, area under the ROC curve; CI, confidence interval; ROC, receiver operating characteristic

**Figure 3.**
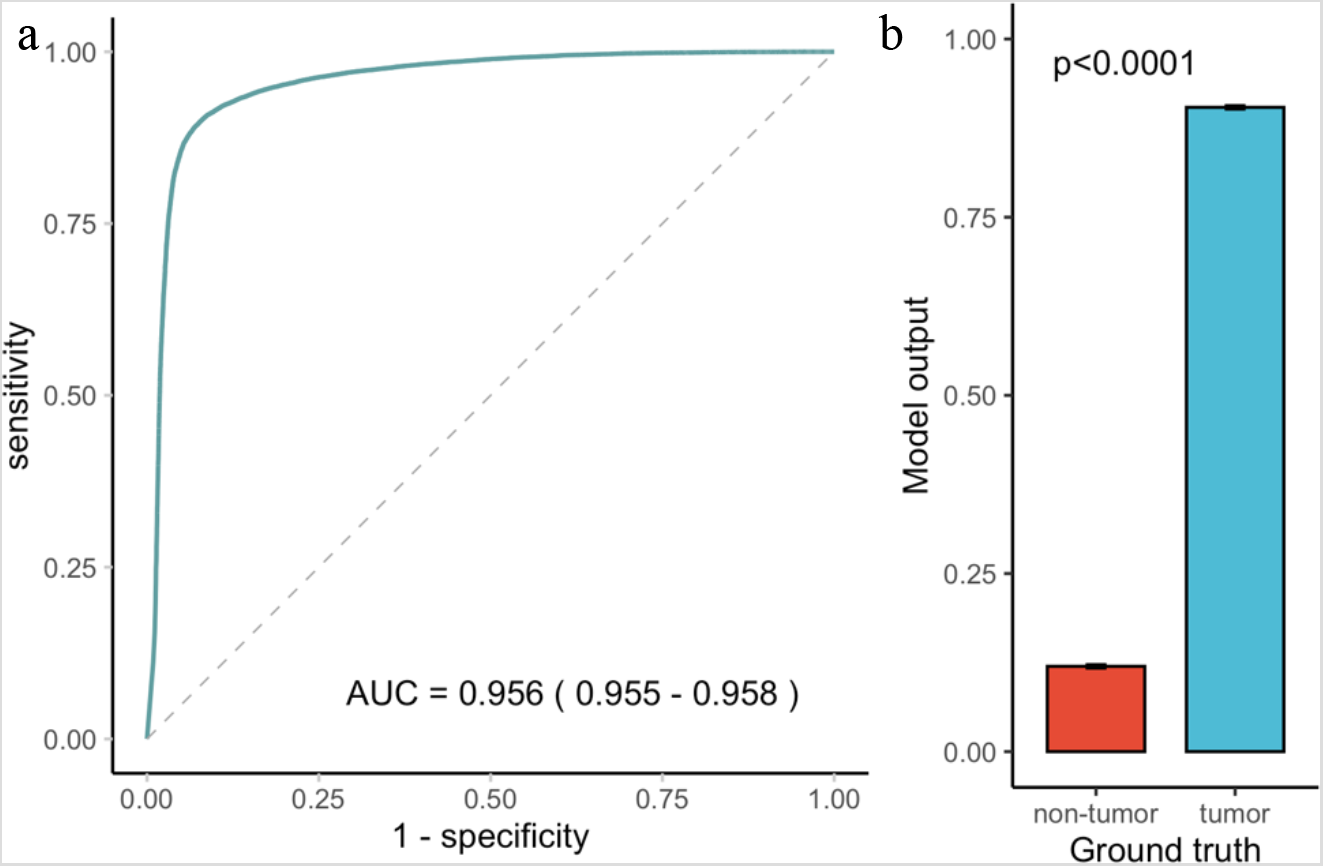
Performance of the tumor tile classification model on the external test set. The AUROC for tumor tile classification was 0.956 (95% CI: 0.955, 0.958) on the external test set (a). Model outputs differed significantly between tiles with a ground truth of tumor versus non-tumor (p-value< 0.0001) (b). *95% CI for AUC is shown in parenthesis in the ROC plot. **Error bars represent 95% CI in the bar chart. The p-value was computed using the Wilcoxon rank sum test. Abbreviations: AUC, area under the ROC curve; CI, confidence interval; ROC, receiver operating characteristic

### Risk score prediction

#### Datasets

To develop a risk score prediction model, we used two datasets: the TCGA-HCC and Stanford-HCC datasets, originating from two independent data sources, the Cancer Genome Atlas(TCGA)-LIHC diagnostic slide collection and the Stanford Department of Pathology slide archive, respectively. The TCGA-HCC was further split into TCGA-HCC development and test datasets.

The TCGA-HCC development dataset (containing the training and validation sets) consisted of 299 patients (median age of 60 years, with an interquartile range (IQR) of 51-68 years, 69% male and 31% female). The frequencies of risk factors for HCC were: 32% for hepatitis B virus infection, 15% for hepatitis C virus infection, 34% for alcohol intake, and 4.9% for NAFLD. The AJCC (8^th^ edition) stage grouping was IA in 2.7%, IB in 41%, II in 29%, IIIA in 20%, IIIB in 5.4%, IVA in 1.0%, and IVB in 0.3% of the patients, respectively. One hundred and fifty-one patients experienced disease recurrence during follow-up (median follow-up time of 12.2 months) (Table 1).

**Table 1:** Patient characteristics for the Stanford-HCC and TCGA-HCC datasets

| Patient characteristic | TCGA-HCC Development Cohort (n=299) | TCGA-HCC Test Cohort (n=53) | Stanford-HCC (n=198) |
| --- | --- | --- | --- |
|  | Training and validation set | Internal test set | External test set |
| **Age (at surgery) (years)** | 60 (51, 68) | 61 (51, 68) | 64 (57, 69) |
| **Gender** |  |  |  |
| Male | 206 (69%) | 33 (62%) | 157 (79%) |
| Female | 93 (31%) | 20 (38%) | 41 (21%) |
| **Hepatitis B virus infection** |  |  |  |
| Negative | 195 (68%) | 33 (67%) | 147 (74%) |
| Positive | 90 (32%) | 16 (33%) | 51 (26%) |
| Unknown | 14 | 4 | 0 |
| **Hepatitis C virus infection** |  |  |  |
| Negative | 243 (85%) | 41 (84%) | 96 (48%) |
| Positive | 42 (15%) | 8 (16%) | 102 (52%) |
| Unknown | 14 | 4 | 0 |
| **Alcohol intake** |  |  |  |
| Negative | 188 (66%) | 30 (61%) | 181 (91%) |
| Positive | 97 (34%) | 19 (39%) | 17 (8.6%) |
| Unknown | 14 | 4 | 0 |
| **Non-alcoholic fatty liver disease** |  |  |  |
| Negative | 271 (95.1%) | 44 (90%) | 184 (93%) |
| Positive | 14 (4.9%) | 5 (10%) | 14 (7.1%) |
| Unknown | 14 | 4 | 0 |
| **AJCC Stage Grouping** |  |  |  |
| IA | 8 (2.7%) | 1 (1.9%) | 44 (22%) |
| IB | 121 (41%) | 24 (46%) | 66 (33%) |
| II | 85 (29%) | 16 (31%) | 68 (34%) |
| IIIA | 60 (20%) | 9 (17%) | 11 (5.6%) |
| IIIB | 16 (5.4%) | 1 (1.9%) | 7 (3.5%) |
| IVA | 3 (1.0%) | 0 (0%) | 2 (1%) |
| IVB | 1 (0.3%) | 1 (1.9%) | 0 (0%) |
| Unknown | 5 | 1 | 0 |
| **Largest tumor diameter (mm)** | 65 (35, 100) | 55 (34, 100) | 30 (18, 50) |
| Unknown | 5 | 0 | 0 |
| **Tumor multifocality** |  |  |  |
| Negative | 207 (69%) | 38 (73%) | 142 (72%) |
| Positive | 91 (31%) | 14 (27%) | 56 (28%) |
| Unknown | 1 | 1 | 0 |
| **Histologic grade** |  |  |  |
| Well-differentiated | 46 (15%) | 5 (9.4%) | 63 (32%) |
| Moderately-differentiated | 162 (54%) | 32 (60%) | 108 (55%) |
| Poorly-differentiated | 89 (30%) | 16 (30%) | 26 (13%) |
| Undifferentiated | 2 (0.7%) | 0 (0%) | 1 (0.5%) |
| **Microvascular invasion** |  |  |  |
| Negative | 196 (67%) | 36 (68%) | 147 (74%) |
| Positive | 95 (33%) | 17 (32%) | 51 (26%) |
| Unknown | 8 | 0 | 0 |
| **Macrovascular invasion** |  |  |  |
| Negative | 271 (93%) | 49 (92%) | 188 (96%) |
| Positive | 21 (7.2%) | 4 (7.5%) | 8 (4.1%) |
| Unknown | 7 | 0 | 0 |
| **Surgical margin status** |  |  |  |
| Negative | 249 (94%) | 45 (88%) | 192 (97%) |
| Positive | 17 (6.4%) | 6 (12%) | 5 (2.5%) |
| Unknown | 33 | 2 | 0 |
| **Fibrosis stage** |  |  |  |
| 0 | 77 (33%) | 13 (30%) | 38 (19%) |
| 1 | 11 (4.8%) | 2 (4.5%) | 13 (6.6%) |
| 2 | 25 (11%) | 4 (9.1%) | 15 (7.6%) |
| 3 | 26 (11%) | 6 (14%) | 13 (6.6%) |
| 4 | 91 (40%) | 19 (43%) | 119 (60%) |
| Unknown | 69 | 9 | 0 |
| **Recurrence** |  |  |  |
| No | 148 (49%) | 28 (53%) | 136 (69%) |
| Yes | 151 (51%) | 25 (47%) | 62 (31%) |
| **Length of follow-up (months)** | 12 (4, 24) | 13 (6, 20) | 25 (9, 48) |
| **Risk score** |  | 0.07 (-0.26, 0.30) | -0.31 (-0.46, -0.15) |

\*Values presented: median (IQR); n (%)

The TCGA-HCC test dataset consisted of 53 patients (median age of 61 years, with an IQR of 51-68 years, 62% male and 38% female). The frequencies of risk factors for HCC were: 33% for hepatitis B virus infection, 16% for hepatitis C virus infection, 39% for alcohol intake, and 10% for NAFLD. The AJCC stage grouping was IA in 1.9%, IB in 46%, II in 31%, IIIA in 17%, IIIB in 1.9%, and IVB in 1.9% of the patients. Twenty-five patients experienced recurrence during follow-up (median follow-up time of 12.7 months) (Table 1). None of the clinicopathologic features were significantly associated with shorter RFI upon univariable Cox regression analysis, while a Batts-Ludwig^22^ fibrosis stage > 2 showed borderline significance (hazard ratio (HR) = 2.7 (95% confidence interval (CI) 0.98, 7.7), p = 0.0543) (Table 2).

**Table 2:** Univariable Cox proportional hazards analysis of the risk of recurrence

| Patient characteristics | TCGA-HCC Test Cohort<br>(n=53) |  | Stanford-HCC (n=198) |  |
| --- | --- | --- | --- | --- |
|  | Internal test set |  | External test set |  |
|  | Hazard ratio (95%<br>CI) | p value | Hazard ratio (95%<br>CI) | p value |
| **Risk score (binarized)** | 6.52 (1.83, 23.2) | 0.0038 | 3.72 (2.17, 6.37) | <0.0001 |
| **Age (at surgery)** |  |  |  |  |
| > 60 | 0.83 (0.38, 1.8) | 0.65 | 1.1 (0.64, 1.8) | 0.77 |
| **Gender** |  |  |  |  |
| Female | 0.99 (0.44, 2.2) | 0.98 | 0.99 (0.53, 1.9) | 0.98 |
| **Hepatitis B virus infection** |  |  |  |  |
| Positive | 0.57 (0.22, 1.5) | 0.25 | 1.1 (0.62, 1.9) | 0.78 |
| **Hepatitis C virus infection** |  |  |  |  |
| Positive | 1.2 (0.4, 3.6) | 0.74 | 0.66 (0.4, 1.1) | 0.11 |
| **Alcohol intake** |  |  |  |  |
| Positive | 0.89 (0.37, 2.2) | 0.80 | 0.19 (0.027, 1.4) | 0.10 |
| **Non-alcoholic fatty liver disease** |  |  |  |  |
| Positive | 1.6 (0.46, 5.4) | 0.48 | 1.8 (0.81, 3.9) | 0.15 |
| **AJCC Stage grouping** |  |  |  |  |
| > II | 1.3 (0.49, 3.6) | 0.58 | 4.4 (2.3, 8.3) | <0.0001 |
| **Largest tumor diameter (mm)** |  |  |  |  |
| > 50 | 1.1 (0.52, 2.5) | 0.74 | 3.5 (2.1, 5.8) | <0.0001 |
| **Tumor multifocality** |  |  |  |  |
| Positive | 1.3 (0.53, 3.4) | 0.53 | 1.1 (0.64, 1.9) | 0.71 |
| **Histologic grade** |  |  |  |  |
| > Moderately-differentiated | 1.1 (0.44, 2.6) | 0.90 | 2.1 (1.2, 3.9) | 0.013 |
| **Microvascular invasion** |  |  |  |  |
| Positive | 1.4 (0.6, 3.1) | 0.46 | 3.9 (2.4, 6.5) | <0.0001 |
| **Macrovascular invasion** |  |  |  |  |
| Positive | 1.6 (0.46, 5.4) | 0.48 | 5.3 (2.1, 1.3) | 0.00043 |
| **Surgical margin** |  |  |  |  |
| Positive | 0.74 (0.22, 2.5) | 0.63 | 6.8 (1.6, 28) | 0.0090 |
| **Fibrosis stage** |  |  |  |  |
| > 2 | 2.7 (0.98, 7.7) | 0.054 | 0.33 (0.2, 0.55) | <0.0001 |

\*CI denotes confidence interval.

The Stanford-HCC dataset consisted of 198 patients (median age of 64 years with an IQR of 57-69 years, 79% male and 21% female). The frequencies of risk factors for HCC were: 26% for hepatitis B virus infection, 52% for hepatitis C virus infection, 8.6% for alcohol intake, and 7.1% for NAFLD. The overall AJCC stage grouping was IA in 22%, IB in 21%, II in 34%, IIIA in 5.6%, IIIB in 3.5%, and IVA in 1.0% of the patients, respectively. Sixty-two patients experienced disease recurrence during follow-up (median follow-up time of 24.9 months) (Table 1). The clinical and pathologic features associated with shorter RFI were AJCC stage grouping > II (HR = 4.4 (95% CI 2.3, 8.3), p< 0.0001), greatest tumor diameter > 5 cm (HR = 3.5 (95% CI 2.1, 5.8), p< 0.0001), histologic grade > moderately differentiated (HR = 2.1 (95% CI 1.2, 3.9), p = 0.0128), presence of microvascular invasion (HR = 3.9 (95% CI 2.4, 6.5), p< 0.0001), presence of macrovascular invasion (HR = 5.3 (95% CI2.1, 13), p< 0.0001), positive surgical margin (HR = 6.8 (95% CI 1.6, 28), p = 0.009), and fibrosis stage > 2 (HR = 0.33 (95% CI 0.2, 0.55), p< 0.0001) using univariable Cox regression analysis (Table 2).

#### HCC-SurvNet performance for RFI prediction

The tumor tile classification model was applied to each tissue-containing image tile in the TCGA-HCC development (n = 299 WSIs) and test (n = 53 WSIs) datasets and the Stanford-HCC dataset (n = 198 WSIs). From each WSI, the 100 tiles with the highest probabilities for the tumor class were selected for input into the subsequent risk score model. Figure 4 shows examples of tiles with probabilities in the top 100 for containing tumor, overlaid onto the original WSI. A MobileNetV2^23^ pre-trained on ImageNet^24^ was modified by replacing the fully-connected layers, and fine-tuned by transfer learning with on-the-fly data augmentation on the tiles from the TCGA-HCC development dataset (n = 307 WSI from 299 patients), where the model input was a 299 × 299 pixel image tile, and the output was a continuous tile-level risk score from the hazard function for RFI. The negative partial log-likelihood of the Cox proportional hazards model was used as a loss function.^14,15^ The model’s performance was evaluated internally on the TCGA-HCC test dataset (n = 53 WSI from 53 patients), and externally on the Stanford-HCC dataset (n = 198 WSI from 198 patients). All tile-level risk scores from a patient were averaged to yield a patient-level risk score.

**Figure 4:**
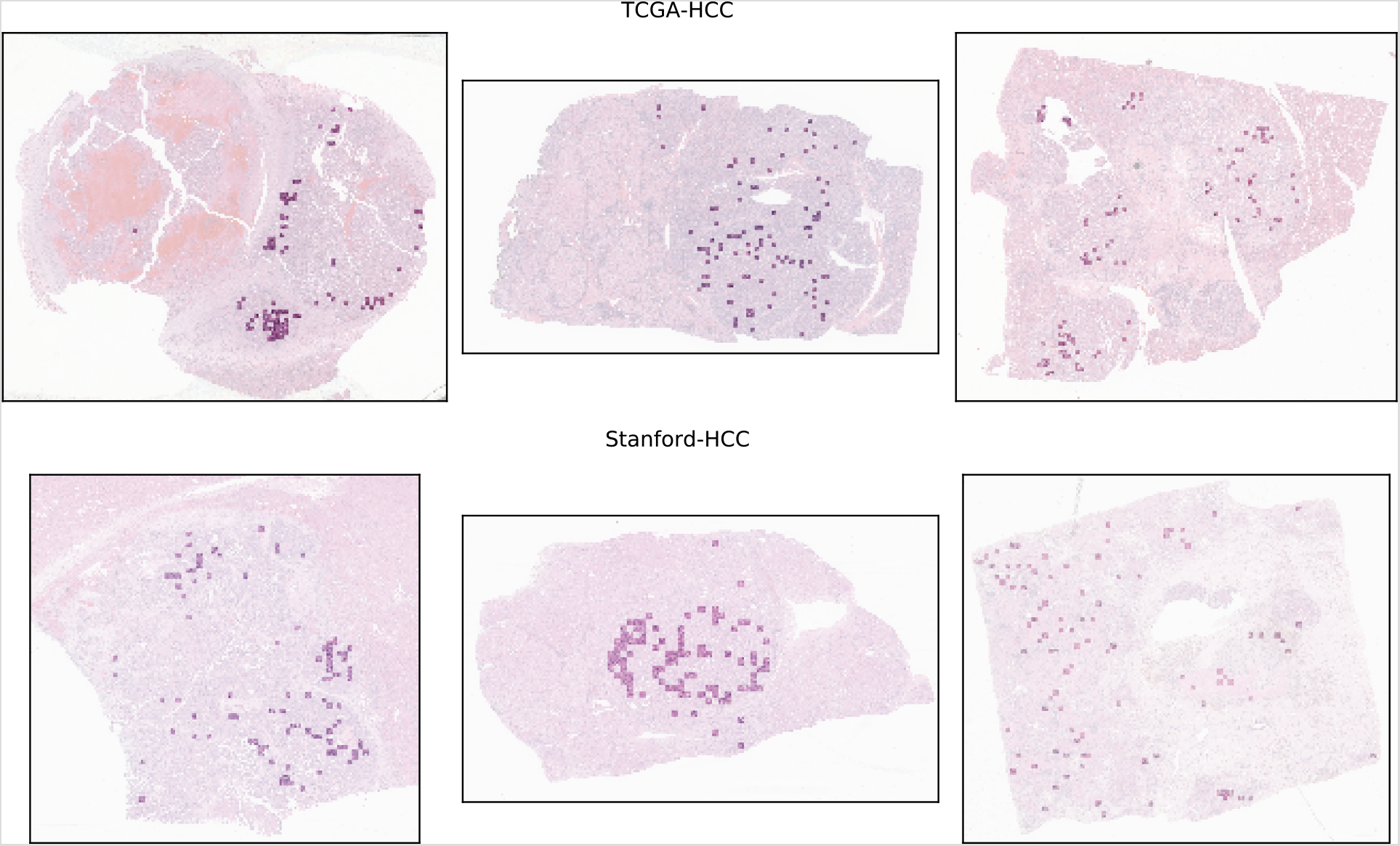
Top 100 tiles selected by the tumor tile classification model. Spatial distribution of the top 100 tiles classified as being tumor tiles by the tumor tile classification model. The top row represents examples from the TCGA-HCC test dataset, and the bottom row represents examples from the Stanford-HCC dataset. The top 100 tiles were subsequently used for development of the survival prediction model. Abbreviation: WSI, whole-slide image

We assessed HCC-SurvNet’s performance using Harrell’s^25^ and Uno’s^26^ concordance indices (c-indices). On the internal test set (TCGA-HCC test dataset, n = 53 patients), Harrell’s and Uno’s c-indices were 0.724 and 0.724, respectively. On the external test set (Stanford-HCC, n = 198 patients), the indices were 0.683 and 0.670, respectively. We observed statistically significant differences in the survival distributions between the low- and high-risk subgroups, as stratified by the risk scores predicted by HCC-SurvNet, on both the internal and external test sets (log-rank p-value: 0.0013 and < 0.0001, respectively) (Figures 5, 6). Histograms of HCC-SurvNet’s risk scores, along with the threshold used for risk group stratification, are shown in Supplementary Figure 1. On univariable Cox proportional hazards analysis, the HCC-SurvNet risk score was a predictor of the RFI, for both the internal (HR = 6.52 (95% CI: 1.83, 23.2), p = 0.0038) and external (HR = 3.72 (95% CI: 2.17, 6.37), p< 0.0001) test sets (Table 2). A continuous linear association between HCC-SurvNet’s risk score and the log relative hazard for RFI was observed by analysis of the internal and external test cohorts by univariable Cox proportional hazards regression with restricted cubic splines (Supplementary Figure 2), validating the use of HCC-SurvNet’s risk score as a linear factor in the Cox analyses.

**Figure 5:**
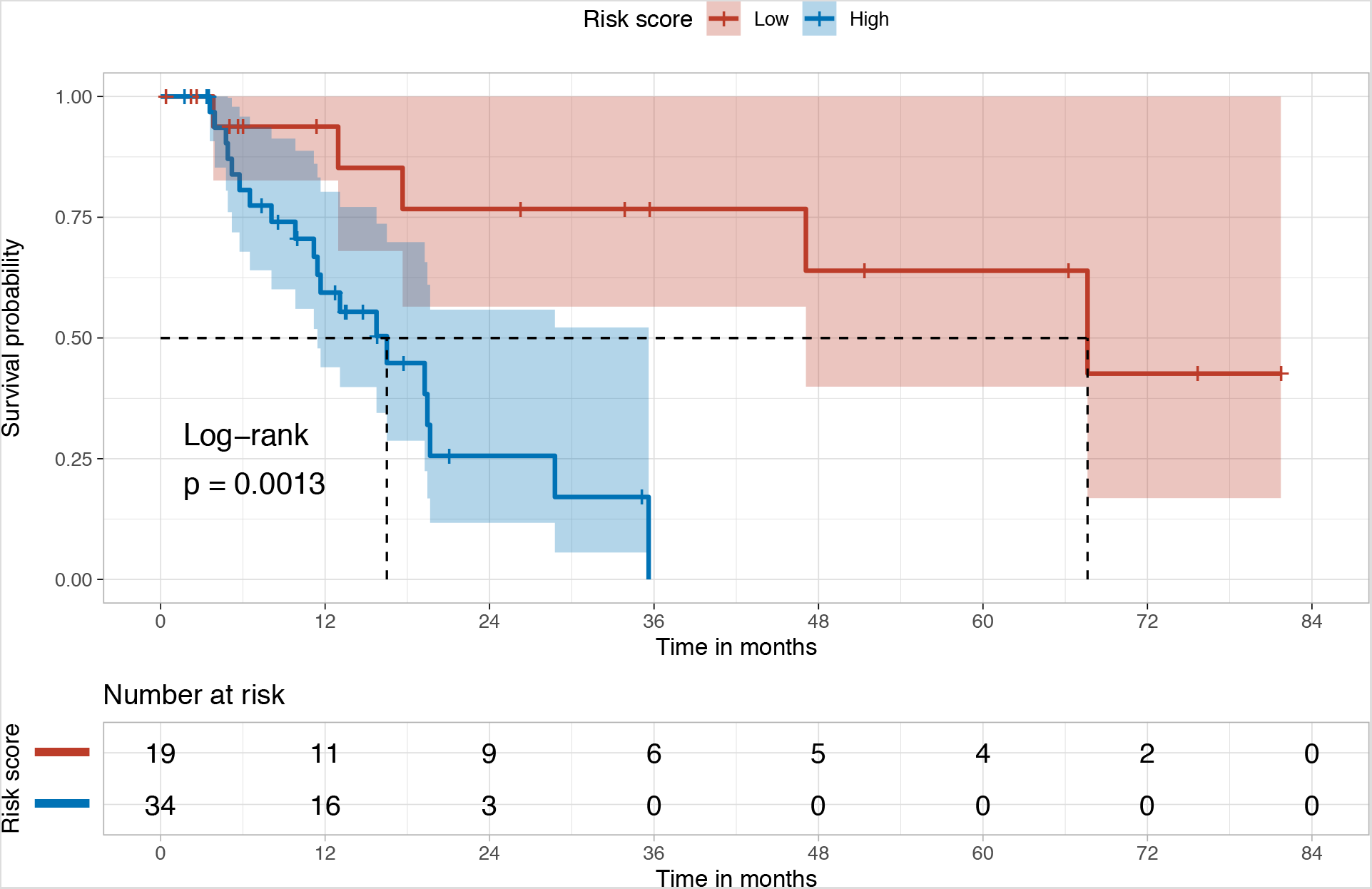
Kaplan-Meier plots for the high- and low-risk subgroups in the internal (TCGA-HCC) test set. The Kaplan-Meier plot shows the difference in the survival distributions for the low- and high-risk subgroups, stratified based on the risk scores predicted by HCC-SurvNet on the internal test set (log-rank p-value = 0.0013).

**Figure 6:**
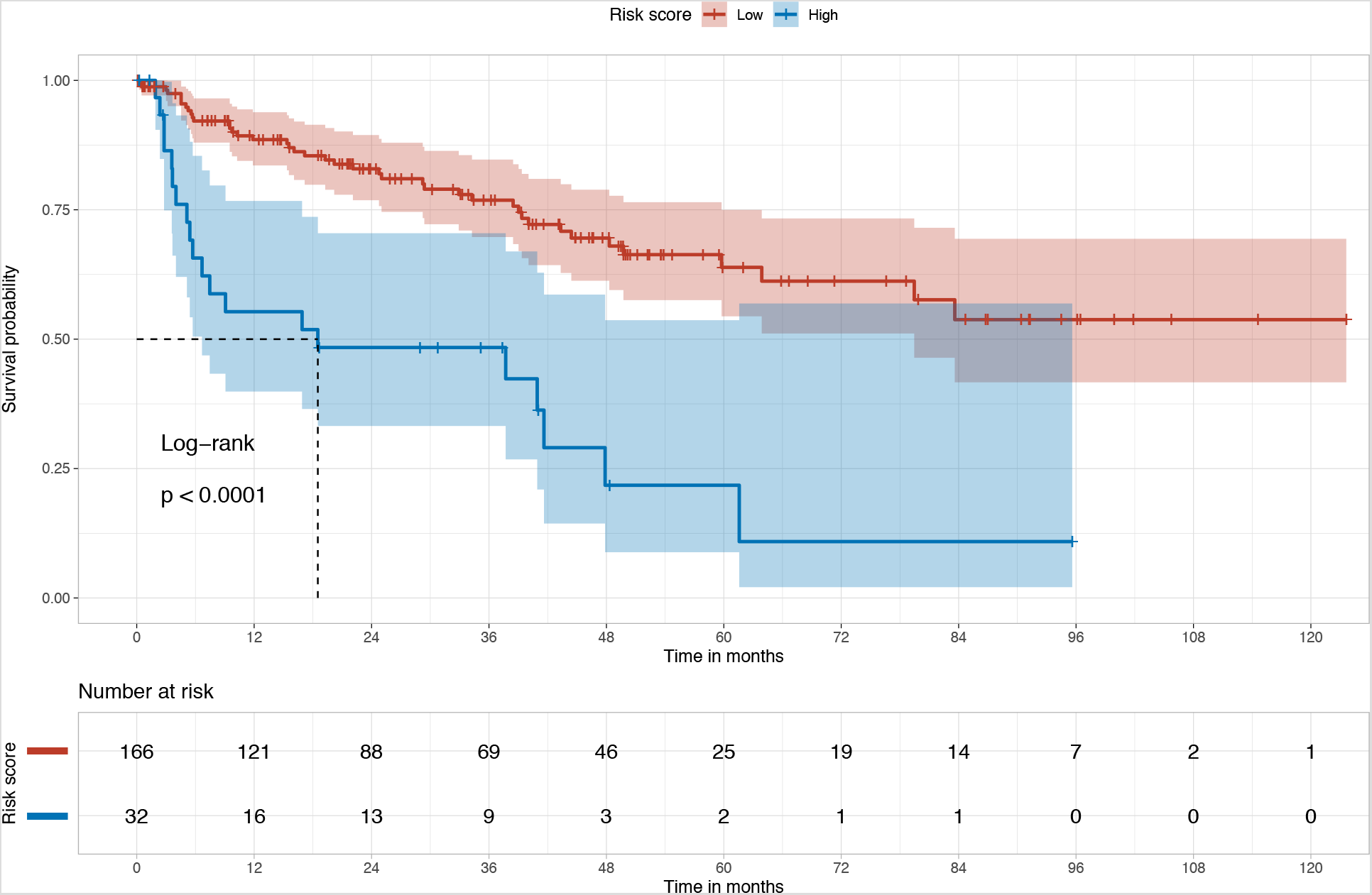
Kaplan-Meier plots for the high- and low-risk subgroups in the external (Stanford-HCC) test set. The Kaplan-Meier plot shows the difference in the survival distributions for the low- and high-risk subgroups, stratified based on the risk scores predicted by HCC-SurvNet on the external test set (log-rank p-value < 0.0001).

On multivariable Cox proportional hazards analysis, HCC-SurvNet’s risk score was an independent predictor of the RFI, for both the internal (HR = 7.44 (95% CI: 1.60, 34.6), p = 0.0105) and external (HR = 2.37 (95% CI: 1.27, 4.43), p = 0.00685) test sets (Table 3). No other clinicopathologic variable was statistically significant on the internal test set. Microvascular invasion (HR = 2.84 (95% CI: 1.61, 5.00), p = 0.000294) and fibrosis stage (HR = 0.501 (95% CI: 0.278, 0.904), p = 0.0217) showed statistical significance on the external test set, along with HCC-SurvNet’s risk score. Schoenfeld’s global test showed p-values greater than 0.05 on both the internal (p = 0.083) and external (p = 0.0702) test sets. On mixed-effect Cox regression analysis with the TCGA institution as a random effect, HCC-SurvNet’s risk score was an independent predictor (p = 0.014), along with the histologic grade (p = 0.014) and macrovascular invasion (p = 0.013). In the external test (Stanford-HCC) cohort, HCC-SurvNet’s risk score was positively associated with the AJCC stage grouping, greatest tumor diameter, and microvascular invasion, and negatively associated with fibrosis stage (Table 4). HCC-SurvNet’s risk score yielded a significantly higher Harrell’s c-index (0.72 for the internal and 0.68 for the external test cohort) than that obtained using the AJCC Stage grouping (0.56 for the internal and 0.60 for the external test cohort), on both the internal and external test cohorts (p = 0.018 and 0.025, respectively).

**Table 3:** Multivariable Cox proportional hazards analysis of the risk of recurrence

| Patient characteristics | TCGA-HCC Test Cohort<br>(n=53) |  | Stanford-HCC (n=198) |  |
| --- | --- | --- | --- | --- |
|  | Internal test set |  | External test set |  |
|  | Hazard ratio (95%<br>CI) | p value | Hazard ratio (95%<br>CI) | p value |
| **Risk score (binarized)** | 7.44 (1.60, 34.6) | 0.011 | 2.37 (1.27, 4.43) | 0.00685 |
| **AJCC Stage grouping** |  |  |  |  |
| > II | 0.30 (0.043, 2.11) | 0.23 | 1.57 (0.63, 3.91) | 0.331 |
| **Largest tumor diameter (mm)** |  |  |  |  |
| > 50 | 1.09 (0.33, 3.60) | 0.89 | 1.32 (0.67, 2.60) | 0.425 |
| **Histologic grade** |  |  |  |  |
| > Moderately-differentiated | 2.82 (0.98, 8.1) | 0.054 | 1.36 (0.69, 2.69) | 0.377 |
| **Microvascular invasion** |  |  |  |  |
| Positive | 1.98 (0.59, 6.64) | 0.27 | 2.84 (1.61, 5.00) | 0.000294 |
| **Macrovascular invasion** |  |  |  |  |
| Positive | 3.76 (0.77, 18.4) | 0.10 | 1.42 (0.42, 4.86) | 0.575 |
| **Surgical margin** |  |  |  |  |
| Positive | 0.51 (0.079, 3.3) | 0.47 | 4.45 (0.84, 23.7) | 0.0797 |
| **Fibrosis stage** |  |  |  |  |
| > 2 | 0.80 (0.20, 3.1) | 0.74 | 0.50 (0.28, 0.90) | 0.0217 |

\*CI denotes confidence interval.

**Table 4:** Association between the HCC-SurvNet risk score and various patient characteristics in the external test (Stanford-HCC) cohort

| Patient characteristics | Stanford-HCC (n=198) |  | Spearman's correlation |  |
| --- | --- | --- | --- | --- |
| | Low-risk group (N = 166) | High-risk group (N = 32) | $\rho$ (95% CI) | p value |
| **Age (at surgery)** | 64 (57, 69) | 64 (57, 69) | -0.017 (-0.15, 0.12) | 0.82 |
| **Gender** |  |  | 0.036 (-0.11, 0.18) | 0.62 |
| Male | 133 (80%) | 24 (75%) |  |  |
| Female | 33 (20%) | 8 (25%) |  |  |
| **Hepatitis B virus infection** |  |  | -0.033 (-0.17, 0.10) | 0.64 |
| Negative | 121 (73%) | 26 (81%) |  |  |
| Positive | 45 (27%) | 6 (19%) |  |  |
| **Hepatitis C virus infection** |  |  | -0.082 (-0.22, 0.063) | 0.25 |
| Negative | 80 (48%) | 16 (50%) |  |  |
| Positive | 86 (52%) | 16 (50%) |  |  |
| **Alcohol intake** |  |  | -0.024 (-0.18, 0.13) | 0.74 |
| Negative | 152 (92%) | 29 (91%) |  |  |
| Positive | 14 (8.4%) | 3 (9.4%) |  |  |
| **Non-alcoholic fatty liver disease** |  |  | -0.0014 (-0.10, 0.092) | 0.99 |
| Negative | 152 (92%) | 32 (100%) |  |  |
| Positive | 14 (8.4%) | 0 (0%) |  |  |
| **AJCC Stage grouping** |  |  | 0.24 (0.086, 0.37) | 0.00079 |
| IA | 43 (26%) | 1 (3.1%) |  |  |
| IB | 53 (32%) | 13 (41%) |  |  |
| II | 57 (34%) | 11 (34%) |  |  |
| IIIA | 7 (4.2%) | 4 (12%) |  |  |
| IIIB | 5 (3.0%) | 2 (6.2%) |  |  |
| IVA | 1 (0.6%) | 1 (3.1%) |  |  |
| IVB | 0 (0%) | 0 (0%) |  |  |
| **Largest tumor diameter (mm)** | 26 (15, 45) | 55 (36, 80) | 0.41 (0.27, 0.53) | <0.0001 |
| **Tumor multifocality** |  |  | -0.10 (-0.24, 0.032) | 0.15 |
| Negative | 188 (71%) | 24 (75%) |  |  |
| Positive | 48 (29%) | 8 (25%) |  |  |
| **Histologic grade** |  |  | 0.14 (-0.0046, 0.27) | 0.058 |
| Well-differentiated | 57 (34%) | 6 (19%) |  |  |
| Moderately-differentiated | 89 (54%) | 19 (59%) |  |  |
| Poorly-differentiated | 19 (11%) | 7 (22%) |  |  |
| Undifferentiated | 1 (0.6%) | 0 (0%) |  |  |
| **Microvascular invasion** |  |  | 0.22 (0.082, 0.35) | 0.0015 |
| Negative | 128 (77%) | 19 (59%) |  |  |
| Positive | 38 (23%) | 13 (41%) |  |  |
| **Macrovascular invasion** |  |  | 0.056 (-0.13, 0.23) | 0.44 |
| Negative | 159 (97%) | 29 (91%) |  |  |
| Positive | 5 (3.0%) | 3 (9.4%) |  |  |
| **Surgical margin status** |  |  | 0.060 (-0.079, 0.18) | 0.40 |
| Negative | 159 (97%) | 29 (91%) |  |  |
| Positive | 5 (3.0%) | 3 (9.4%) |  |  |
| Unknown | 2 | 0 |  |  |
| **Fibrosis stage** |  |  | -0.36 (-0.48, -0.22) | <0.0001 |
| 0 | 29 (17%) | 9 (28%) |  |  |
| 1 | 8 (4.8%) | 5 (16%) |  |  |
| 2 | 12 (7.2%) | 3 (9.4%) |  |  |
| 3 | 9 (5.4%) | 4 (12%) |  |  |
| 4 | 108 (65%) | 11 (34%) |  |  |

\*Values presented: median (IQR); n (%)

## Discussion

Building upon recent advances in deep learning, we have developed a system for predicting RFI after curative-intent surgical resection in patients with HCC, directly from H&E-stained FFPE WSI. The system outputs an RFI risk score by first applying a deep CNN to automatically detect tumor-containing tiles. Then, a second model outputs a continuous risk score based on analysis of the top 100 tumor-containing tiles from each WSI. In the internal and external test cohorts, we observed statistically significant differences in the survival distributions between the low- and high-risk subgroups, as stratified by the risk score predicted by the system. The results of multivariable analyses indicate that the HCC-SurvNet risk score could help supplement established clinicopathologic predictors of RFI, thereby improving recurrence risk stratification.

In the present study, HCC-SurvNet significantly outperformed the standard AJCC/UICC staging system in predicting the post-surgical HCC recurrence risk. Shim et al.^12^ reported the performance of a prognostic nomogram for recurrence prediction after curative liver resection in HCC patients, which yielded a c-index of 0.66 for 2-year recurrence on an independent validation cohort. Although a direct comparison with the performance of their nomogram is not possible, as their patient cohort was different from ours, our risk score appears to have a performance that is on par with, or slightly better than, the prognostic nomogram.

Advances in deep learning, and other forms of machine learning, have led to the identification of histomorphologic features informative of disease outcomes, and prior works have applied these methods to automated outcome prediction.^14–21^ The automatic extraction of such features directly from WSI has the potential to add value to current treatment planning paradigms by increasing both the accuracy of prognostic risk stratification and the objectivity and reproducibility of biomarker assessment. Mobadersany et al.^14^ and Zhu et al.^15^ previously applied convolutional neural networks to survival prediction directly from histopathologic images, by integrating the negative partial log-likelihood into the model as a loss function, which enables the model to output a value that can be regarded as a prognostic risk score. However, in these prior studies, representative tiles were manually identified for input into the deep learning models. This requirement for manual tile selection, even during inference, makes such models less practical for widespread clinical deployment. In this work, we present a system which automatically selects representative image tiles, which should increase the ease of deployment in clinical settings.

Saillard et al.^21^ were the first to apply deep learning to digital H&E WSI to predict overall survival after resection in HCC patients. On their external test set (342 WSI from the TCGA), their models yielded c-indices of 0.68 and 0.70 for overall survival prediction. We were unable to perform a direct comparison with their models, as the outcomes and datasets used were different, but our c-index on the external test set of 0.68 appears comparable to that reported for their model. In their study, they applied a CNN pre-trained on ImageNet as a fixed feature extractor. The features extracted were optimized for natural, rather than histopathologic, images, suggesting that there might be further potential for improving prediction performance by optimizing feature extraction for histopathologic images.^27^ To leverage the full capacity of our HCC-SurvNet deep learning system, we fine-tuned all of the models’ parameters, including those for feature extraction (i.e. the convolutional blocks), with histopathologic images. Whereas models to date have focused on predicting overall survival, ours focused on the recurrence-free interval, as the intent was to aid refinement of treatment strategies by providing a risk score that was specific for HCC recurrence and/or HCC-related mortality after curative-intent surgical resection.

A specific strength of our study was the review and confirmation of all clinicopathologic variables in the TCGA-HCC cohort and re-coding of older edition AJCC classifications to the latest 8^th^ edition classification. Previous other studies^19^ have also developed models for the prediction of overall survival in post-surgical HCC patients, also by using clinicopathologic data from the TCGA. However, use of TCGA clinicopathologic data presents some significant limitations, which are often overlooked. These include the fact that the AJCC TNM classifications used across cases in the TCGA-LIHC dataset range from the 4^th^ through the 7^th^ editions, resulting in inconsistency in the meaning of the pathologic T, N, and M categories across different patients resected during different time periods. In addition, the pathology reports in TCGA-LIHC came from different institutions with wide variation in the reporting of pathologic features. Therefore, prior to use of TCGA data, standardization, in particular, of all pathologic variables, as performed in this study, is necessary. As the TCGA data were collected from 35 different institutions, each with different H&E staining and digitization protocols, we constructed a mixed-effect Cox model to account for potential intraclass correlations present between WSI originating from the same institution. After taking the originating institution into account as a random effect, we found that HCC-SurvNet’s risk score remained an independent predictor of recurrence-free interval, along with the histologic grade and the presence of macrovascular invasion.

A limitation of our study was that the dataset used to externally evaluate HCC-SurvNet’s performance was restricted to cases from a single institution. Due to limitations in the datasets that were available to us, we chose to reserve the more heterogeneous, multi-institutional TCGAHCC dataset for HCC-SurvNet model development, with the intention of capturing histomorphologic features informative of HCC recurrence which were robust to inter-institutional variations in H&E staining and scanning protocols. With further development and validation on larger, more diverse datasets, we hope that risk scores produced by HCC-SurvNet, as well as other similar deep learning-based models, might one day offer clinical value as a supplement to currently-established clinicopathologic predictors of recurrence and survival.

Another limitation was the black-box nature of deep learning systems. To gain insights into model interpretability, we assessed the associations between HCC-SurvNet’s risk score and different patient characteristics in the external test (Stanford-HCC) cohort. The HCC-SurvNet risk score was significantly associated with several well-recognized prognostic factors, including the AJCC stage grouping, largest tumor diameter, microvascular invasion, and Batts-Ludwig fibrosis stage. In addition, the independent contribution of the HCC-SurvNet risk score to recurrence-free interval prediction, when analyzed together with other known clinicopathologic variables in the multivariable Cox regression, suggests that HCC-SurvNet was able to extract some as-yet unrecognized histomorphologic features informative of recurrence, which might have biological significance and correlate with other important outcomes, such as response to adjuvant treatment. It remains for future studies to explore the additional potential of deep learning for prognostication and treatment response prediction in HCC, and other malignancies.

In conclusion, we have shown that a deep learning-based cancer recurrence risk score extracted from routine H&E WSI of primary surgical resections for HCC independently predicts the RFI, and significantly outperforms the most commonly-used standard AJCC/UICC stage grouping. With further validation on larger, more diverse datasets, such a risk score could augment current methods for predicting the risk of HCC recurrence after primary surgical resection, thereby assisting clinicians in tailoring post-surgical management.

## Methods

### Patient population

A total of 250 primary hepatic resection specimens (n = 250 patients) from surgeries performed at our institution between January 1, 2009 and December 31, 2017, with glass slides available for retrieval from the departmental slide archive, were included in the dataset. Prior to digitization, the time to recurrence after surgical resection, as well as patient demographic information (age at surgical resection, gender, and alcohol intake), and clinicopathologic variables (history of hepatitis B and C viral infection, non-alcoholic fatty liver disease (NAFLD), HCC multi-nodularity, macro- and micro-vascular invasion, largest tumor diameter, histologic World Health Organization grade,^28^ Batts-Ludwig^22^ fibrosis stage, surgical margin status, and AJCC (8^th^ edition) stage^3^) were collected for each case by review of the electronic health records by trained physicians at Stanford University Medical Center (J.S. and A. S.). Forty-seven patients were excluded because their resections were performed for recurrent HCC, two were excluded because of lack of follow-up data after surgical resection, and three were excluded due to the presence of comorbidities known to have contributed to the patients’ deaths. This process narrowed the final number of study patients down to 198. From each of these 198 patients, a representative tumor H&E slide (the one containing the highest grade of tumor in the specimen) was digitized at high resolution (40x objective magnification, 0.25 micrometers per pixel) on an Aperio AT2 scanner (Leica Biosystems, Nussloch, Germany), to generate a WSI in the SVS file format. This dataset (n = 198 WSI, from 198 unique patients), referred to as the Stanford-HCC dataset, was used for external evaluation of the risk score prediction model. From the excluded patient pool (not included in Stanford-HCC), 36 patients were randomly selected, and a representative tumor H&E slide from each patient was digitized using the exact same method as described above, yielding a dataset with 36 WSI from 36 patients, referred to as the Stanford-HCCDET dataset. This dataset was used to develop a model for automatically detecting tumor-containing tiles in a WSI (“DET” stands for “detection”). Use of all patient material and data was approved by the Stanford University Institutional Review Board, with waived informed consent.

In addition to the Stanford-HCC and Stanford-HCCDET datasets, a publicly-available dataset of 379 FFPE diagnostic WSI from 365 unique patients in the TCGA-LIHC diagnostic slide collection were downloaded via the GDC Data Portal^29^ and used to develop the risk score prediction model for this study. The same patient demographics, clinicopathologic variables, and RFI as collected for Stanford-HCC were obtained through review of the accompanying metadata and pathology reports downloaded from the GDC Data Portal and the previously-published Integrated TCGA Pan-Cancer Clinical Data Resource by Liu et al.^30^ RFI was defined as the period from the date of surgery until the date of the first occurrence of a new tumor event, which included progression of HCC, locoregional recurrence, distant metastasis, new primary tumor, or death with tumor.^30^ Patients who were alive without these events, or who died without tumor, were censored.^31^ The event time was the shortest period from the date of surgery to the date of an event. The censored time was the period from the date of surgery to the date of last contact with the patient or the date of death without HCC. Given multiple changes to the AJCC classification over the time period during which these specimens were collected (resulting in differences in the pathologic staging criteria across different editions of the AJCC), a reference pathologist trained in the interpretation of hepatobiliary pathology (J.S.) reviewed the WSI and the downloaded pathology reports, in order to re-stage all of the patients based on the most current AJCC (8th edition) classification.^3^ WSI scanned at 20x base magnification were excluded (n = 10 WSI, from 4 patients). One patient (n = 1 WSI) with missing RFI was excluded. Seven patients (n = 7 WSIs) with mixed HCC–cholangiocarcinomas and one patient (n = 1 WSI) with an angiomyolipoma were excluded from the dataset. The final dataset (n = 360 WSI, from 352 patients), referred to as the TCGA-HCC dataset, contained patients from 35 institutions, each with potentially different staining and scanning protocols. The TCGA-HCC dataset was randomly split into the development cohort (n = 299 patients: n = 247 patients for training and n = 52 patients for validation) and internal test cohort (n = 53 patients), with no patient overlap between the splits.

### WSI image preprocessing

First, tissue segmentation (i.e. tissue separation from white background) of the WSI was performed by applying a combination of filters. Second, the WSI were tiled into image patches with a size of 1024 × 1024 pixels, at a resolution of 40x (0.25 µm/pixel). Only the tiles containing an overall tissue percentage of > 80% of the total surface area within each tile were saved in PNG format. Lastly, the Vahadane method^32^ was used for stain normalization, to convert all image tiles to a reference color space. All tiles were subsequently resized to 299 × 299 pixels and used for the downstream analyses.

### Tumor tile classification

All tumor regions in each WSI in the Stanford-HCCDET dataset were manually annotated by the reference pathologist (J.S.) at 10x magnification, using Aperio ImageScope (Leica Biosystems, Nussloch, Germany). Tiles containing both tumor and normal tissue were excluded from model development and evaluation. Using these ground-truth annotated WSI, we developed a CNN for automatically classifying an image tile into either the tumor or non-tumor class, where the model input was a 299 × 299 pixel image tile in PNG format, and the output was a probability for each class. The particular CNN architecture, PathCNN, which was originally proposed by Bilaloglu et al,^33^ was trained and tested using the Stanford-HCCDET (n = 128,222 tiles from 36 WSI) dataset, with 78% of WSI used for training, 11% used for validation, and 11% used as an internal test set, with no patient overlap between any of these three sets). We used leaky ReLU^34^ with negative slope 0.01 as the non-linearity. The dropout probability was set at 0.1. The trainable parameters were initialized using a Xavier weight initialization scheme,^35^ and updated using an Adam optimization method^36^ with an initial learning rate of 0.001. We applied stepwise learning rate decay with a step size of 7 and gamma of 0.1. The number of epochs was set at 25, with a mini-batch size of 32. A binary cross entropy loss function was applied. Input images were normalized by ((image – 0.5) / 0.5) before passing them to the model. We augmented the training data by randomly introducing positional transforms: a horizontal flip and a rotation of 0°, 90°, 180° or 270° degrees. Additionally, we randomly adjusted the hue, brightness, contrast, and saturation of the image. We used validation accuracy to select the final model. The final optimized tumor versus non-tumor tile classifier was externally tested on 30 WSI (n = 82,532 tiles) randomly sampled from the TCGA-HCC dataset. Of note, there was no patient overlap between the Stanford-HCCDET and Stanford-HCC datasets, where the latter was used in the downstream development of the risk score prediction model. The tumor tile classification model was subsequently applied to each tissue-containing image tile in the Stanford-HCC (n = 198 WSIs) and TCGA-HCC (n = 360 WSIs) datasets. From each WSI, the 100 tiles with the highest probabilities for the tumor class were selected for input into the subsequent survival analysis. The value of 100 was chosen arbitrarily in order to incorporate enough representative tiles, taking into account morphologic tumor heterogeneity in the WSI (Figure 4). Additional details on the model’s development are described in the Supplementary Methods.

### HCC-SurvNet Development

The top 100 tiles selected by the tumor detector were used for the development of the risk score model for RFI, which consisted of a MobileNetV2^23^ pre-trained on ImageNet,^24^ modified by replacing the fully-connected layers, and fine-tuned by transfer learning with on-the-fly data augmentation on the tiles from the TCGA-HCC development dataset (n = 307 WSI, n = 299 patients), where the model input was a 299 × 299 pixel image tile in PNG format, and the output was a continuous tile-level risk score from the hazard function for RFI. The dropout probability in the replaced fully-connected classification layers was set at 0.7. The trainable parameters were fine-tuned using an AdamW optimization method^37^ with an initial learning rate of 0.001. The number of epochs was set at 30, with a mini-batch size of 80. The negative partial log-likelihood of the Cox proportional hazards model was used as a loss function.^14,15^ Input images were normalized by ((image – mean) / standard deviation), where the mean and standard statistics were calculated for the ImageNet dataset before passing them to the model. We augmented the training data by randomly introducing positional transforms: a horizontal flip and a rotation of 0°, 90°, 180° or 270° degrees. Additionally, we randomly adjusted the hue, brightness, contrast, and saturation of the image. We used validation loss to select the final model. The model’s performance was evaluated internally on the TCGA-HCC test dataset, and externally on the Stanford-HCC dataset. All tile-level risk scores from a patient were averaged to yield a patient-level risk score. An overall framework for the system, referred to as HCC-SurvNet, is shown in Figure 1, with additional model development details described in the Supplementary Methods.

### Hardware and software

The PyTorch Python package (version 1.1.0)^38^ was used for model development. OpenSlide (version 3.4.1)^39^ was used to read WSI in the SVS format. Image preprocessing was performed on a High-Performance Computing (HPC) cluster operated by the Stanford Research Computing Center (Sherlock cluster: https://www.sherlock.stanford.edu/). Model development and evaluation were performed on a workstation with two GeForce RTX 2080 Ti (NVIDIA, Santa Clara, CA) graphics processing units, a Core i9-9820X (10 cores, 3.3 GHz) central processing unit (Intel, Santa Clara, CA), and 128 GB of random-access memory.

### Statistical analysis

We summarized our study population with descriptive statistics, including the median and IQR for continuous variables, and the proportion for categorical variables. The performance of the tumor tile classification model was assessed using the overall accuracy and AUROC. Model outputs for tiles with a ground truth of tumor were compared with those for tiles with a ground truth of non-tumor, using the Wilcoxon rank sum test. We evaluated the performance of the risk score model using Harrell’s^25^ and Uno’s^26^ c-indices, which indicate better prediction when their values approach one. Each patient was stratified into one of two subgroups (high-risk and low-risk), based on their patient-level risk score. The median risk score on the validation set from TCGA-HCC was used as the threshold for patient stratification (Supplementary Figure 1). Kaplan-Meier analysis was performed, and a log-rank test was used to compare the survival distributions between the subgroups. Univariable and multivariable Cox proportional hazards models were used to assess the relationship between independent variables and RFI. The independent variables included HCC-SurvNet’s risk score, age at surgical resection, gender, AJCC stage grouping, largest tumor diameter, tumor multifocality, histologic tumor grade, microvascular invasion, macrovascular invasion, surgical margin status, fibrosis stage, and history of Hepatitis B, Hepatitis C, alcohol intake, and non-alcoholic fatty liver disease. Of these, variables with univariable p-values of less than 0.1 on either the internal or external test sets were selected for inclusion in the multivariable analysis. The proportional hazards assumption was checked using Schoenfeld’s global test. To demonstrate the non-linear relationship between HCC-SurvNet’s risk score and the log relative hazard for RFI, univariable Cox proportional hazards regression analysis with restricted cubic splines (3 knots) was performed. To account for potential intraclass correlation among WSI prepared and scanned at the same institution within the TCGA cohort, a mixed-effect Cox regression model was constructed using the institution as a random effect. Spearman’s correlation coefficients were computed to gain insight into associations between the HCC-SurvNet risk score and different patient characteristics in the external test (Stanford-HCC) cohort. Harrell’s c-index was compared between HCC-SurvNet’s risk score and the standard AJCC staging system, using a paired t-test.

A two-tailed alpha level of 0.05 was used for statistical significance. All statistical analyses were performed using Python (v3.6.4, Python Software Foundation, https://www.python.org/) with the lifelines (v0.24.0) and scikit-survival (v0.11) packages, as well as R (v3.6.3, R Foundation for Statistical Computing, http://www.R-project.org/) with the survival (v3.1.12), coxme (v2.2.16), pROC (v1.16.2), and rms (v5.1.4) packages.

## Data Availability

All whole-slide-images for the TCGA cohort are publicly available at https://portal.gdc.cancer.gov/. The Stanford whole-slide images are not publicly available, in accordance with institutional requirements governing human subject privacy protection.

https://github.com/RubinLab/HCCSurvNet

## Acknowledgments

This work was funded by the Stanford Departments of Pathology and Biomedical Data Science, through a Stanford Clinical Data Science Fellowship to R.Y. Additional computational infrastructure was provided by the Stanford Research Computing Center. We would also like to thank Dr. Lu Tian, Stanford Department of Biomedical Data Science, for helpful initial conversations regarding analysis planning.

## Author Contributions

R.Y., D.L.R., and J.S. conceived and designed the study; R.Y. and J.S. performed the literature search. R.Y., A.S. and J.S. performed the data collection; R.Y. performed the model development and performance evaluation; R.Y. and J.L. performed the statistical analyses; R.Y. drafted the manuscript; D.L.R. and J.S. supervised the study; all authors participated in the critical revision and approval of the manuscript.

## Code availability

All source code is available under an open-source license at: https://github.com/RubinLab/HCCSurvNet

## Supplementary Figures

**Supplementary Figure 1:**
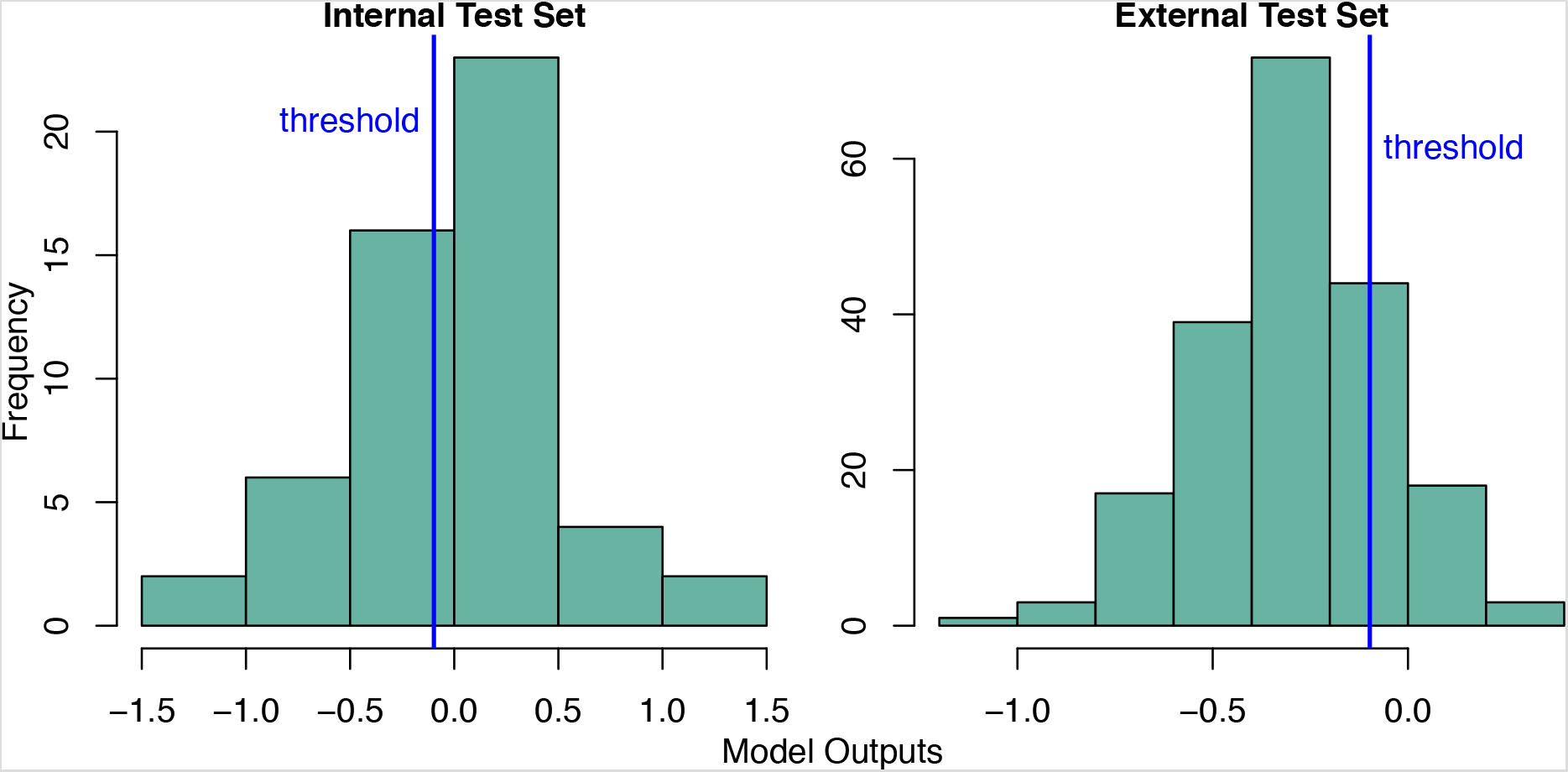
Histograms of HCC-SurvNet Risk Scores. The histograms show the distributions of HCC-SurvNet’s risk scores within the internal (left) and external (external) test sets separately. A threshold used for patient stratification into low-and high-risk groups, which was determined on the validation set from TCGA-HCC, is visualized as a blue vertical line (threshold = −0.0978).

**Supplementary Figure 2:**
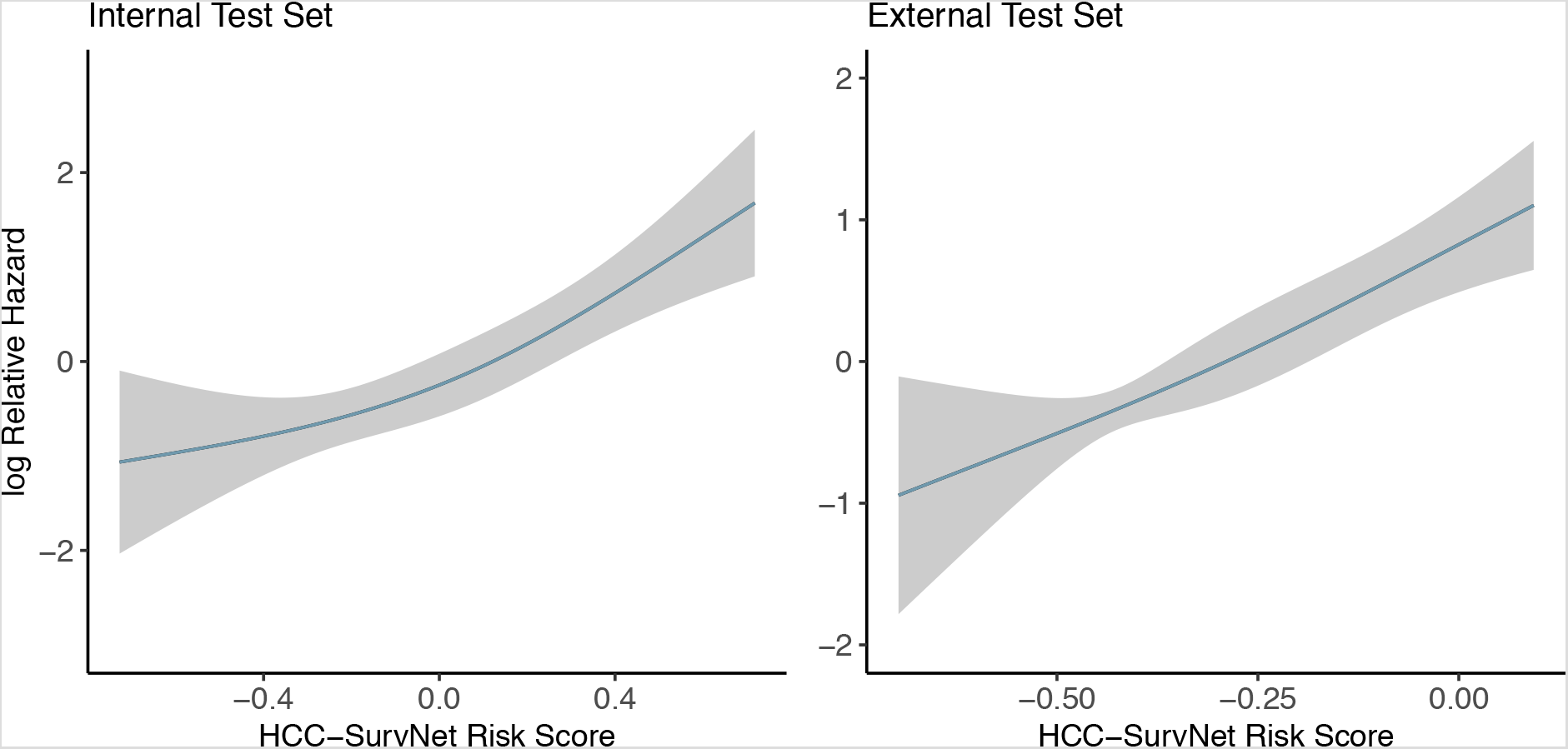
Univariable Cox Proportional Hazards Regression Analysis with Restricted Cubic Splines. A continuous linear association between HCC-SurvNet’s risk score and the log relative hazard for RFI was observed upon analysis of the internal test set (left), and even more significantly for the external test set (right). The blue line represents the fitted line of the association between HCC-SurvNet’s risk score and the log relative hazard for RFI; the shaded region represents the 95% CI. Abbreviations: CI, confidence interval; RFI, recurrence-free interval

